# Galectin-3 as a potential prognostic biomarker of severe COVID-19 in SARS-CoV-2 infected patients

**DOI:** 10.1101/2021.02.07.21251281

**Authors:** Eduardo Cervantes-Alvarez, Nathaly Limon-de la Rosa, Moisés Salgado-de la Mora, Paola Valdez-Sandoval, Mildred Palacios-Jiménez, Fátima Rodriguez-Alvarez, Brenda I. Vera-Maldonado, Eduardo Aguirre-Aguilar, Juan Manuel Escobar-Valderrama, Jorge Alanis-Mendizabal, Osvely Méndez-Guerrero, Farid Tejeda-Dominguez, Jiram Torres-Ruíz, Diana Gómez-Martín, Kathryn L Colborn, David Kershenobich, Christene A Huang, Nalu Navarro-Alvarez

**Affiliations:** Instituto Nacional de Ciencias Médicas y Nutrición Salvador Zubirán, Department of Gastroenterology, Mexico City, Mexico; Instituto Nacional de Ciencias Médicas y Nutrición Salvador Zubirán, Department of Internal Medicine, Mexico City, Mexico; Instituto Nacional de Ciencias Médicas y Nutrición Salvador Zubirán, Department of Immunology and Rheumatology, Mexico City, Mexico; Universidad Panamericana School of Medicine, Campus México, Mexico City; PECEM, Facultad de Medicina, Universidad Nacional Autónoma de México, Mexico City, Mexico; Universidad Veracruzana, Veracruz, Mexico; Department of Surgery, University of Colorado Anschutz Medical Campus, Denver, CO

**Author notes:** Corresponding Author: **Nalu Navarro-Alvarez, MD PhD**, Instituto Nacional de Ciencias Médicas y Nutrición Salvador Zubirán Vasco de Quiroga #15, Tlalpan, **Email:**. Equally contributed (Eduardo Cervantes-Alvarez, Nathaly Limon-de la Rosa).

## Abstract

**BACKGROUND:** Prognostic biomarkers are needed to identify patients at high-risk for severe COVID-19. Galectin-3 is known to drive neutrophil infiltration and release of pro-inflammatory cytokines contributing to airway inflammation.

**METHODS:** In this prospective cohort, we assessed galectin-3 levels in 156 hospitalized patients with confirmed COVID-19. COVID-19 patients were diagnosed as either *critical* (>50% lung damage) or *moderate* (<50% of lung damage) based on computerized tomography. Patients who required invasive mechanical ventilation (IMV) and/or died during hospitalization were categorized as having a *severe outcome*, and a *non-severe outcome* if they were discharged and none of the former occurred.

**RESULTS:** Elevated serum galectin-3 was significantly higher in critical patients compared to moderate ones (35.91 ± 19.37 ng/mL vs. 25 ± 14.85 ng/mL, p<0.0001). Patients who progressed to a *severe outcome* including IMV and/or in-hospital death, presented higher galectin-3 levels (41.17 ng/mL [IQR 29.71 – 52.25] vs. 23.76 ng/mL [IQR 15.78 – 33.97] compared to those of a non-severe outcome, p<0.0001). Galectin-3 discriminated well between those with severe and non-severe outcome, with an AUC of 0.75 (95% CI 0.67 – 0.84, p<0.0001) and was found to be an independent predictor of severe outcome regardless of the percentage of lung involvement. Additionally, the combination of galectin-3, CRP and albumin, significantly improved its individual predicting ability with an AUC 0.84 (95% CI 0.77 – 0.91, p<0.0001).

**CONCLUSION:** Circulating galectin-3 levels can be used to predict severe outcomes in COVID-19 patients, including the requirement of mechanical ventilation and/or death, regardless of the initial severity of the disease.

## INTRODUCTION

Coronavirus disease 2019 (COVID-19) caused by severe acute respiratory syndrome coronavirus 2 (SARS-CoV-2) infection has afflicted tens of millions of people in a worldwide pandemic, straining health care systems across the world (1, 2). Prognostic biomarkers that can identify high-risk patients are needed to improve clinical management and allow appropriate allocation of healthcare resources. Moreover, the lack of current curative therapies emphasizes the need to get a better understanding of the pathophysiological process behind SARS-CoV-2 infection and its long-term consequences for the development of targeted therapeutic strategies.

Severe COVID-19 is associated with a systemic hyperinflammatory response characterized by high levels of circulating cytokines and chemokines (3) and substantial lung infiltration of innate immune cells (4) that can lead to acute respiratory distress syndrome (ARDS), multi-organ failure and death (5, 6). Among the inflammatory cytokines are those associated with the activation of monocyte/macrophages such as Interleukin 6 (IL-6), Tumor necrosis factor (TNF), and the CC-chemokine ligand 2 (CCL2) (3, 6, 7).

Studies have shown that those inflammatory cytokines contribute to the recruitment of additional inflammatory cells that not only aggravate the lung damage, but also lead to pulmonary fibrosis (8, 9). Subsets of M2 macrophages expressing profibrogenic genes have been found in the bronchoalveolar lavage of COVID-19 patients (4), reflecting that the pathological process of SARS-CoV-2 infection not only involves an acute inflammatory response in the lungs, but is also associated with fibrotic complications (10).

Galectin-3 is a 29-35 kDa ß-galactoside binding lectin first identified in macrophages (11). It plays an important role as a driver and amplifier of the pro-inflammatory response by promoting the release of several cytokines including IL-6 and TNF-α (12), which are some of the major cytokines present in severe COVID-19 patients (3). High levels of galectin-3 are known to drive neutrophil infiltration and the release of pro-inflammatory cytokines, contributing to acute airway inflammation (13-15). In addition, studies have shown that endogenous galectin-3 can enhance the effects of viral infection by promoting host inflammatory responses (16, 17).

Galectin-3 is increasingly recognized as a potentially important diagnostic or prognostic biomarker for a variety of inflammatory and fibrotic diseases (18, 19),(20). Galectin-3 has been implicated in the development of organ fibrosis and is found to be highly upregulated in the injured lung, particularly in patients with idiopathic pulmonary fibrosis (21, 22). Elevated circulating galectin-3 levels were previously reported to be associated with disease severity and mortality in ARDS patients (23).

Inflammation and fibrosis are key contributing mechanisms to the progression of severe COVID-19 and the development of its long-term consequences (3),(10, 24). Given the known proinflammatory and profibrotic roles of galectin-3, we measured circulating galectin-3 levels in COVID-19 patients to assess its prognostic value.

In this prospective study we followed 156 patients with confirmed COVID-19 from admission to discharge or death. We hypothesized that levels of serum galectin-3 upon hospital admission could identify patients at high-risk of progressing to a severe COVID-19 outcome resulting in invasive mechanical ventilation (IMV) and/or death. Additionally, galectin-3 levels were correlated with clinical and inflammatory laboratory markers. COVID-19 patients were diagnosed as either critical (>50% lung damage) or moderate (<50% of lung damage) based on computerized tomography (CT). We found that elevated serum galectin-3 was significantly higher in critical patients compared to moderate ones and was found to be an independent predictor of severe outcome regardless of the percentage of lung involvement. Results from this study indicate that galectin-3 may be a useful prognostic biomarker in COVID-19 patients to provide early identification of patients at high risk of severe illness and to provide guidance on resource allocation. Further studies are warranted to understand the potential pathophysiological role of galectin-3 in COVID-19 disease progression and the development of targeted therapies.

## RESULTS

### Demographic, clinical and laboratory characteristics

A total of 156 patients with real-time reverse transcriptase-polymerase chain reaction (RT-PCR)-confirmed SARS-CoV-2 infection and CT findings were enrolled in the study, of which 107 (68.6%) were male and 49 (31.4%) female. According to the CT scan images, pulmonary affection was greater than 50% in 94 patients (critical patients) and less than 50% in 62 (moderate patients). Critical patients were significantly older (54.89 ± 11.79 vs. 48.21 ± 14.30, p<0.01) and the majority were male (71 [66.4%] vs. 36 [33.6%], p=0.02). The principal comorbidities among our cohort were obesity (44.2%), hypertension (30.1%) and diabetes (21.2%), all of which had been diagnosed prior to hospital admission. Alcohol consumption was higher in critical patients (p<0.04).

Critical patients had a significantly longer hospital stay (9 [6 – 18] vs. 6 [4 – 8], p<0.0001) and more often required IMV (44 [46.8%] vs. 7 [11.3%], p<0.0001), both of which reflected a superior need of hospital care. The overall mortality rate was 13.5% (n=21), and critical patients had the highest mortality rate (18 [19.1%] vs. 3 [4 .8%], p=0.02). A total of 54 (34.6%) patients developed a severe outcome, including IMV and/or in-hospital death, and the majority of these patients were critical (47 [50%] vs. 7 [11.3 %], p<0.0001) (**Table 1A**).

**Table 1.**
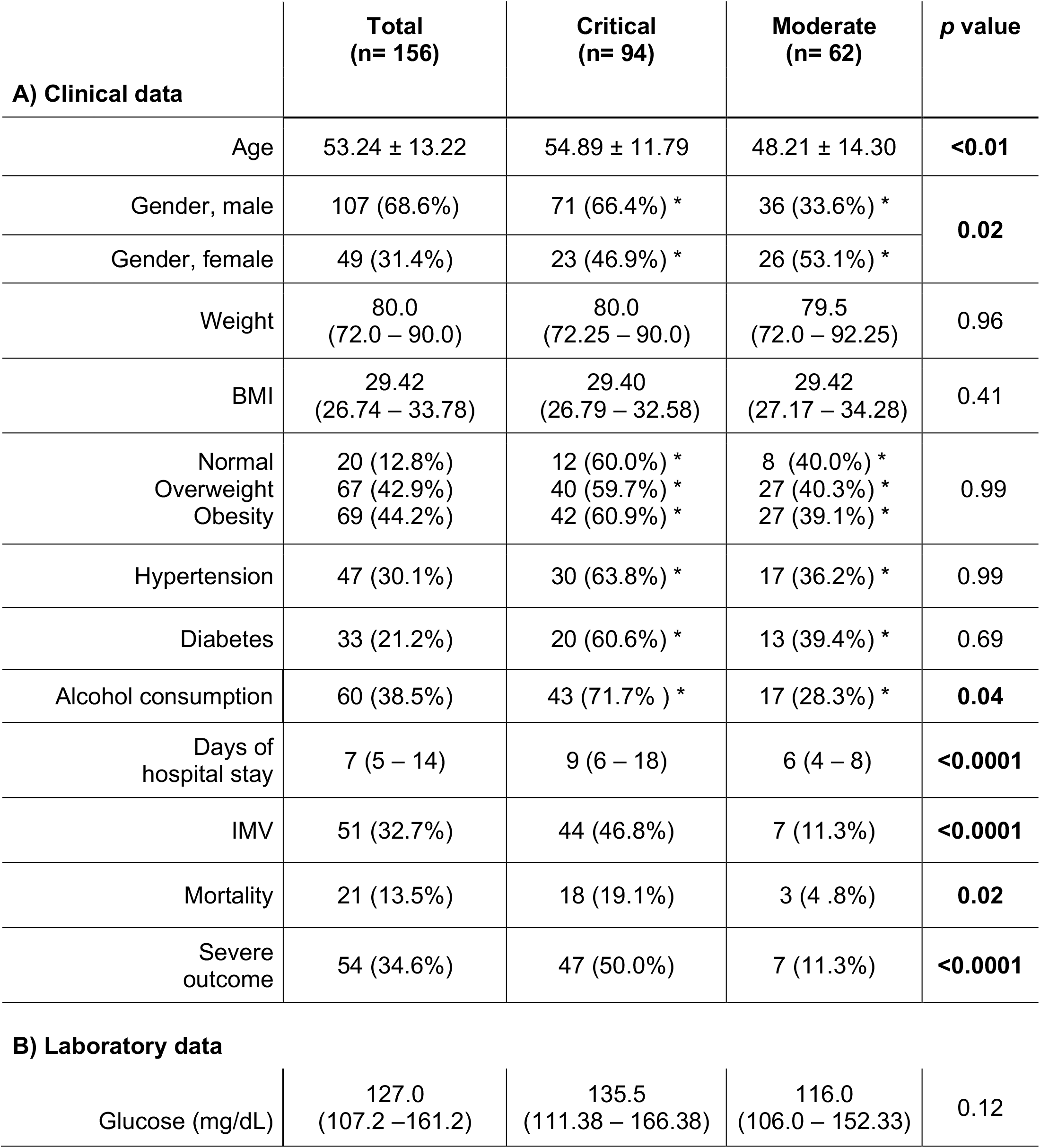
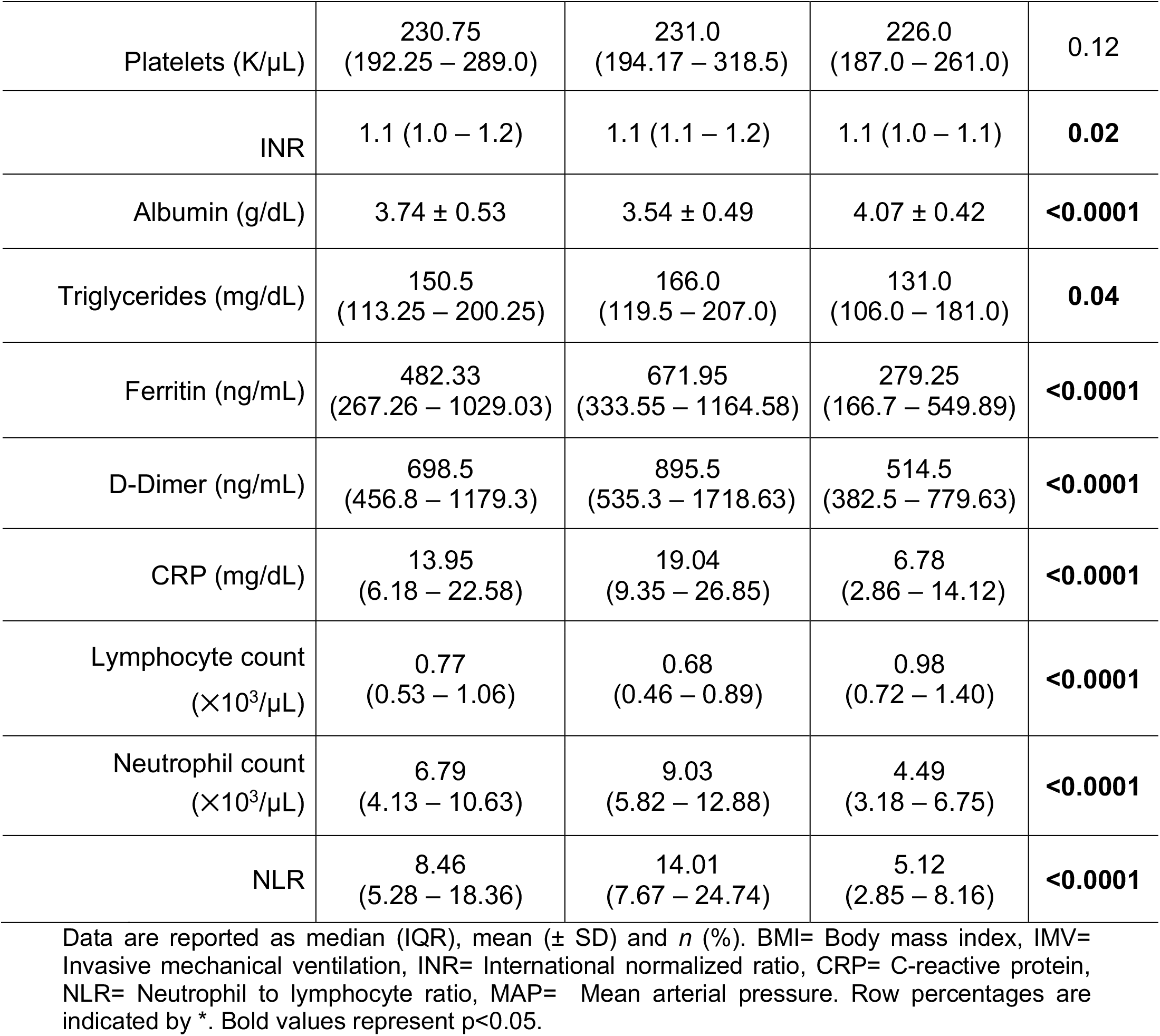
Demographic, clinical and laboratory characteristics.

|  | **Total**<br>**(n= 156)** | **Critical**<br>**(n= 94)** | **Moderate**<br>**(n= 62)** | **p value** |
| --- | --- | --- | --- | --- |
| **A) Clinical data** |  |  |  |  |
| Age | 53.24 ± 13.22 | 54.89 ± 11.79 | 48.21 ± 14.30 | **<0.01** |
| Gender, male | 107 (68.6%) | 71 (66.4%) * | 36 (33.6%) * | **0.02** |
| Gender, female | 49 (31.4%) | 23 (46.9%) * | 26 (53.1%) * |  |
| Weight | 80.0<br>(72.0 – 90.0) | 80.0<br>(72.25 – 90.0) | 79.5<br>(72.0 – 92.25) | 0.96 |
| BMI | 29.42<br>(26.74 – 33.78) | 29.40<br>(26.79 – 32.58) | 29.42<br>(27.17 – 34.28) | 0.41 |
| Normal | 20 (12.8%) | 12 (60.0%) * | 8 (40.0%) * | 0.99 |
| Overweight | 67 (42.9%) | 40 (59.7%) * | 27 (40.3%) * |  |
| Obesity | 69 (44.2%) | 42 (60.9%) * | 27 (39.1%) * |  |
| Hypertension | 47 (30.1%) | 30 (63.8%) * | 17 (36.2%) * | 0.99 |
| Diabetes | 33 (21.2%) | 20 (60.6%) * | 13 (39.4%) * | 0.69 |
| Alcohol consumption | 60 (38.5%) | 43 (71.7%) * | 17 (28.3%) * | **0.04** |
| Days of hospital stay | 7 (5 – 14) | 9 (6 – 18) | 6 (4 – 8) | **<0.0001** |
| IMV | 51 (32.7%) | 44 (46.8%) | 7 (11.3%) | **<0.0001** |
| Mortality | 21 (13.5%) | 18 (19.1%) | 3 (4 .8%) | **0.02** |
| Severe outcome | 54 (34.6%) | 47 (50.0%) | 7 (11.3%) | **<0.0001** |
| **B) Laboratory data** |  |  |  |  |
| Glucose (mg/dL) | 127.0<br>(107.2 – 161.2) | 135.5<br>(111.38 – 166.38) | 116.0<br>(106.0 – 152.33) | 0.12 |
| Platelets (K/μL) | 230.75<br>(192.25 – 289.0) | 231.0<br>(194.17 – 318.5) | 226.0<br>(187.0 – 261.0) | 0.12 |
| INR | 1.1 (1.0 – 1.2) | 1.1 (1.1 – 1.2) | 1.1 (1.0 – 1.1) | **0.02** |
| Albumin (g/dL) | 3.74 ± 0.53 | 3.54 ± 0.49 | 4.07 ± 0.42 | **<0.0001** |
| Triglycerides (mg/dL) | 150.5<br>(113.25 – 200.25) | 166.0<br>(119.5 – 207.0) | 131.0<br>(106.0 – 181.0) | **0.04** |
| Ferritin (ng/mL) | 482.33<br>(267.26 – 1029.03) | 671.95<br>(333.55 – 1164.58) | 279.25<br>(166.7 – 549.89) | **<0.0001** |
| D-Dimer (ng/mL) | 698.5<br>(456.8 – 1179.3) | 895.5<br>(535.3 – 1718.63) | 514.5<br>(382.5 – 779.63) | **<0.0001** |
| CRP (mg/dL) | 13.95<br>(6.18 – 22.58) | 19.04<br>(9.35 – 26.85) | 6.78<br>(2.86 – 14.12) | **<0.0001** |
| Lymphocyte count<br>($\times 10^3/\mu$L) | 0.77<br>(0.53 – 1.06) | 0.68<br>(0.46 – 0.89) | 0.98<br>(0.72 – 1.40) | **<0.0001** |
| Neutrophil count<br>($\times 10^3/\mu$L) | 6.79<br>(4.13 – 10.63) | 9.03<br>(5.82 – 12.88) | 4.49<br>(3.18 – 6.75) | **<0.0001** |
| NLR | 8.46<br>(5.28 – 18.36) | 14.01<br>(7.67 – 24.74) | 5.12<br>(2.85 – 8.16) | **<0.0001** |

Data are reported as median (IQR), mean ( ± SD) and *n* (%). BMI= Body mass index, IMV= Invasive mechanical ventilation, INR= International normalized ratio, CRP= C-reactive protein, NLR= Neutrophil to lymphocyte ratio, MAP= Mean arterial pressure. Row percentages are indicated by \*. Bold values represent $p < 0.05$ .

Laboratory data obtained upon hospital admission are summarized in **Table 1B**. While glucose levels were slightly elevated in critical patients (135.5 mg/dL [111.38 – 166.38] vs. 116.0 mg/dL [106.0 – 152.33], p=0.12), the platelet count was similar in both groups (231.0 K/uL [194.17 – 318.5] vs. 226.0 K/uL [187.0 – 261.0], p=0.12) for which no significant difference was found in either laboratory parameter between those diagnosed as critical or moderate based on CT findings. However, albumin and triglycerides were statistically different between critical and moderate patients. Albumin levels were significantly lower (3.54 ± 0.49 g/dL vs. 4.07 ± 0.42, p<0.0001) in critical patients compared to moderate patients while triglycerides were significantly higher (166.0 mg/dL [119.5 – 207.0] vs. 131.0 mg/dL [106.0 – 181.0], p=0.04).

The inflammatory markers ferritin, D-dimer and C-reactive protein (CRP) were all significantly elevated (p<0.0001) upon hospital admission in critical patients (671.95 ng/mL [333.55– 1164.58] vs. 279.25 ng/mL [166.7 – 549.89]; 895.5 ng/mL [535.3 – 1718.63] vs. 514.5 ng/mL [382.5 – 779.63]; and 19.04 mg/dL [9.35 – 26.85] vs. 6.78 mg/dL [2.86 – 14.12], respectively), presumably reflecting a greater inflammatory state due to more severe lung damage. The total lymphocyte count was significantly lower in critical patients (0.68 ×10^3^/µL [0.46 – 0.89] vs. 0.98 ×10^3^/µL [0.72 – 1.40], p<0.0001). In contrast, neutrophil count was significantly higher in patients characterized as critical based on CT upon admission (9.03 ×10^3^/µL [5.82 – 12.88] vs. 4.49 ×10^3^/µL [3.18 vs. 6.75], p<0.0001). Both lymphocyte and neutrophil findings are reflected in the NLR ratio, which was higher in critical patients (14.01 [7.67 – 24.74] vs. 5.12 [2.85 – 8.16], p<0.0001).

### Galectin-3 serum levels are higher in COVID-19 patients with a severe outcome

To explore the possible role of galectin-3 as a biomarker of severity, circulating levels were measured in sera from COVID-19 patients using a commercial enzyme-linked immunosorbent assay (ELISA). We found that COVID-19 patients upon hospital admission had significantly elevated circulating levels of galectin-3 when compared to age-matched pre-pandemic healthy subjects (28.77 ng/mL [IQR 17.52 – 42.04] vs. 9.65 ng/mL [IQR 8.27 – 14.71], p<0.0001) (**Supplementary table 1**; **Figure 1a**). Critical patients had significantly higher levels of galectin-3 when compared to moderate patients (35.91 ± 19.37 ng/mL vs. 25.0 ± 14.85 ng/mL, p<0.001) (**Figure 1b)**. Patients who developed a severe outcome, including IMV and/or in-hospital death, had significantly higher galectin-3 levels than those with a non-severe outcome, (41.17 ng/mL [IQR 29.71 – 52.25] vs. 23.76 ng/mL [IQR 15.78 – 33.97], p<0.0001) (**Figure 1c**). When outcomes were analyzed according to the initial disease state, galectin-3 levels were significantly higher in those with a severe outcome (39.21 ng/mL [IQR 28.01 – 50.85] vs. 26.60 ng/mL [IQR 20.22 – 42.59, p<0.01 in critical patients and 50.03 ± 11.88 vs. 21.81 ± 11.91 ng/mL, p<0.0001, in moderate patients) regardless of their initial diagnose by CT (**Figure 2a**). Galectin-3 not only predicted an adverse outcome in critical patients but was able to even more accurately identify patients within the moderate group likely to progress in disease severity (AUC 0.66 vs. 0.95) (**Figure 2b - c**). We next analyzed CRP levels between critical and moderate patients according to outcome. Similar to findings with galectin-3, CRP was found to be higher in those with severe outcome regardless of the initial disease state (21.90 ± 9.56 mg/dL vs. 15.25 ± 9.50, p<0.01 in critical patients and 17.80 mg/dL [IQR 15.36 – 21.30] vs. 6.49 mg/dL [IQR 2.70 – 12.66], p<0.01, in moderate patients) (**Figure 2d**). When comparing the predictive ability of CRP for a severe outcome in critical and moderate patients, CRP performed very similar to galectin-3 when the critical group was analyzed (AUC=0.70 vs. AUC=0.66, respectively) (**Figure 2e**). However, galectin-3 had additional power and significance in predicting severe outcomes in the moderate group compared to CRP (AUC=0.95, p<0.0001, and AUC=0.87, p<0.01, respectively) (**Figure 2f**).

**Fig. 1:**
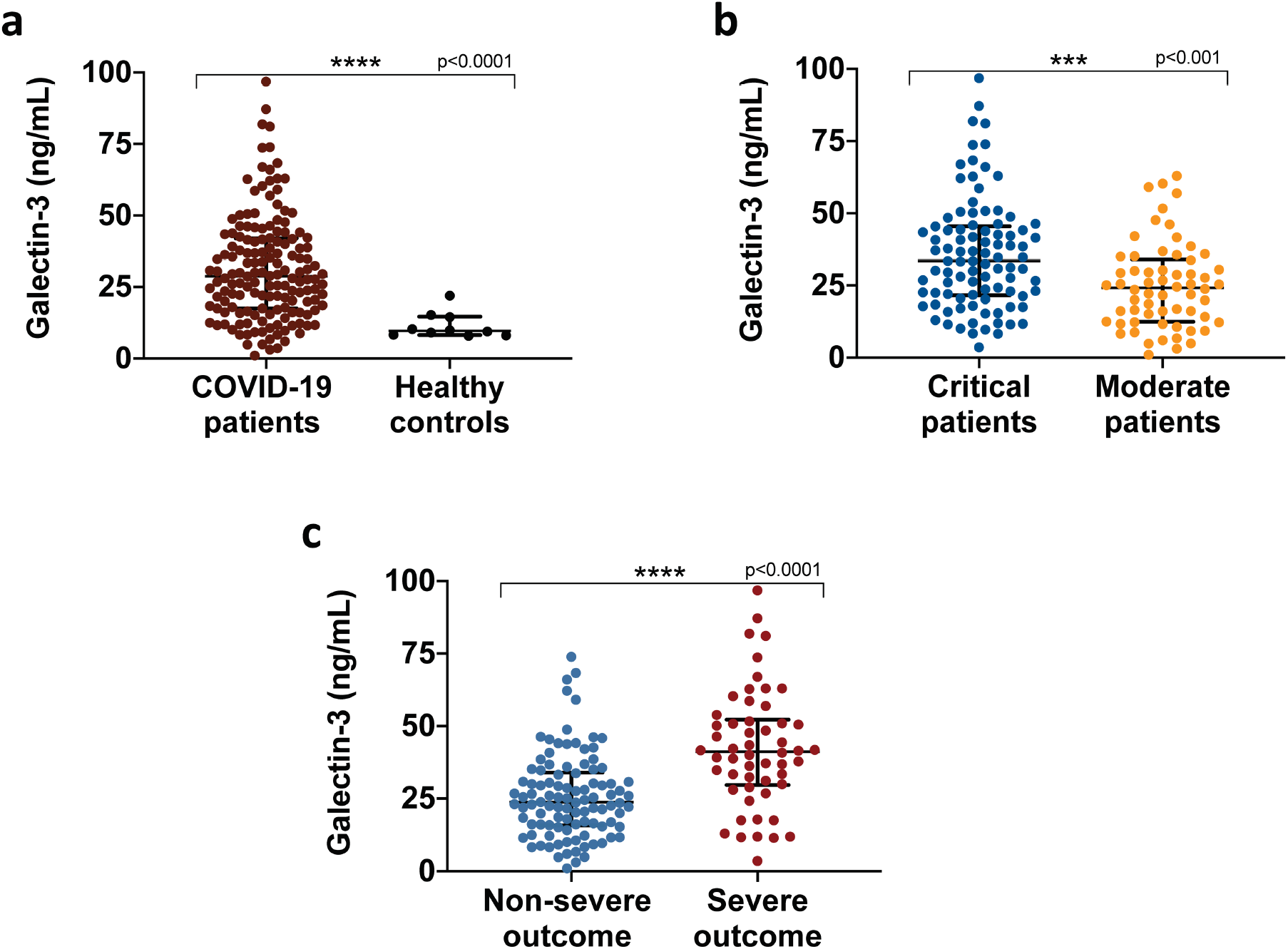
Galectin-3 serum levels in COVID-19 patients. **a**, Galectin-3 circulating levels upon hospital admission of COVID-19 patients (n=156) and age-matched healthy pre-pandemic controls (n=10). **b**, Galectin-3 is associated with COVID-19 severity, critical patients (n=94) presented significantly higher levels than moderate patients (n=62). **c**, Severe outcomes in COVID-19 patients were associated with elevated levels of galectin-3. Data in **a** and **c** are shown as median with IQR, data in **b** as mean ± SD. \*\*\**p* < 0.001, \*\*\*\**p* < 0.0001; two-tailed Mann-Whitney U test or two-tailed t-test. Samples were assessed in duplicate in ELISA assays.

**Fig. 2:**
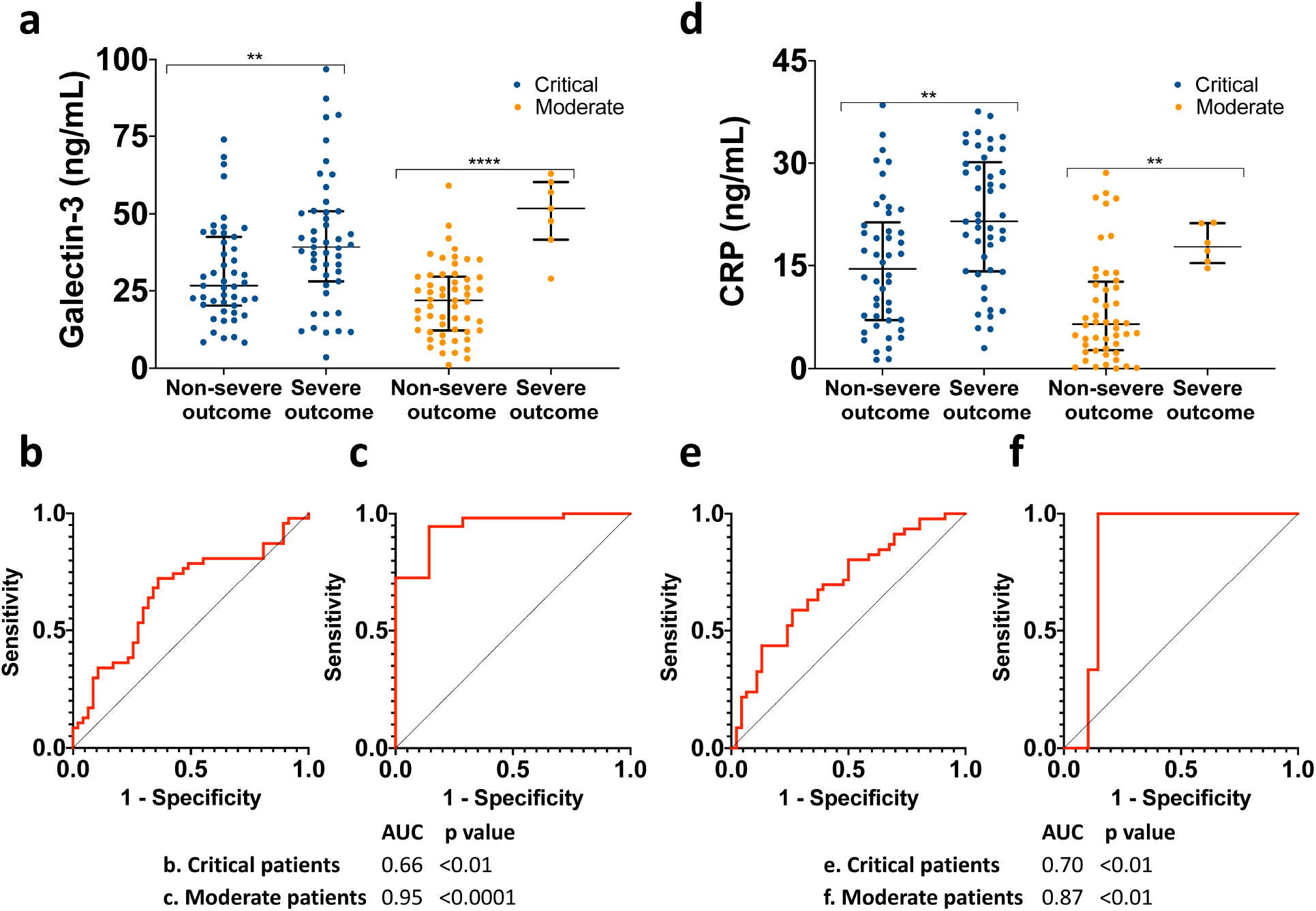
Galectin-3 and CRP as predictors of severe outcome in moderate and critical COVID-19 patients. **a**, significantly higher galectin-3 levels associate with severe outcome in both critical and moderate groups **b**,**c**, Receiver-operating characteristic (ROC) curves show that galectin-3 has a significant predictive power for severe outcome in critical (b) and moderate patients (c). **d**, Similarly, C-reactive protein (CRP) levels are elevated in critical and moderate patients with severe outcome. **e, f**, Predictive power is also found with CRP when ROC curves for severe outcome are plotted in both groups. Data in **a** and **d** are shown as median with IQR. \*\**p* < 0.01, \*\*\*\**p* < 0.0001; two-tailed Mann-Whitney U test.

### Galectin-3 correlates with other inflammatory biomarkers

Given that elevated galectin-3 levels were found in patients who progressed to a severe outcome, and considering its participation in the inflammatory response, correlations with inflammatory parameters previously studied in COVID-19 using the spearman correlation coefficient, in accordance with the non-normal distribution of the data were carried out. We found that galectin-3 correlates positively with CRP (r=0.42, p<0.0001), neutrophil count (r=0.39, p<0.0001), D-dimer (r=0.19, p=0.02), ferritin (r=0.30, p<0.001), LDH (r=0.36, p<0.0001); whereas a negative correlation was found with albumin (r= −0.35, p<0.0001) (**Figure 3a – f**).

**Fig. 3:**
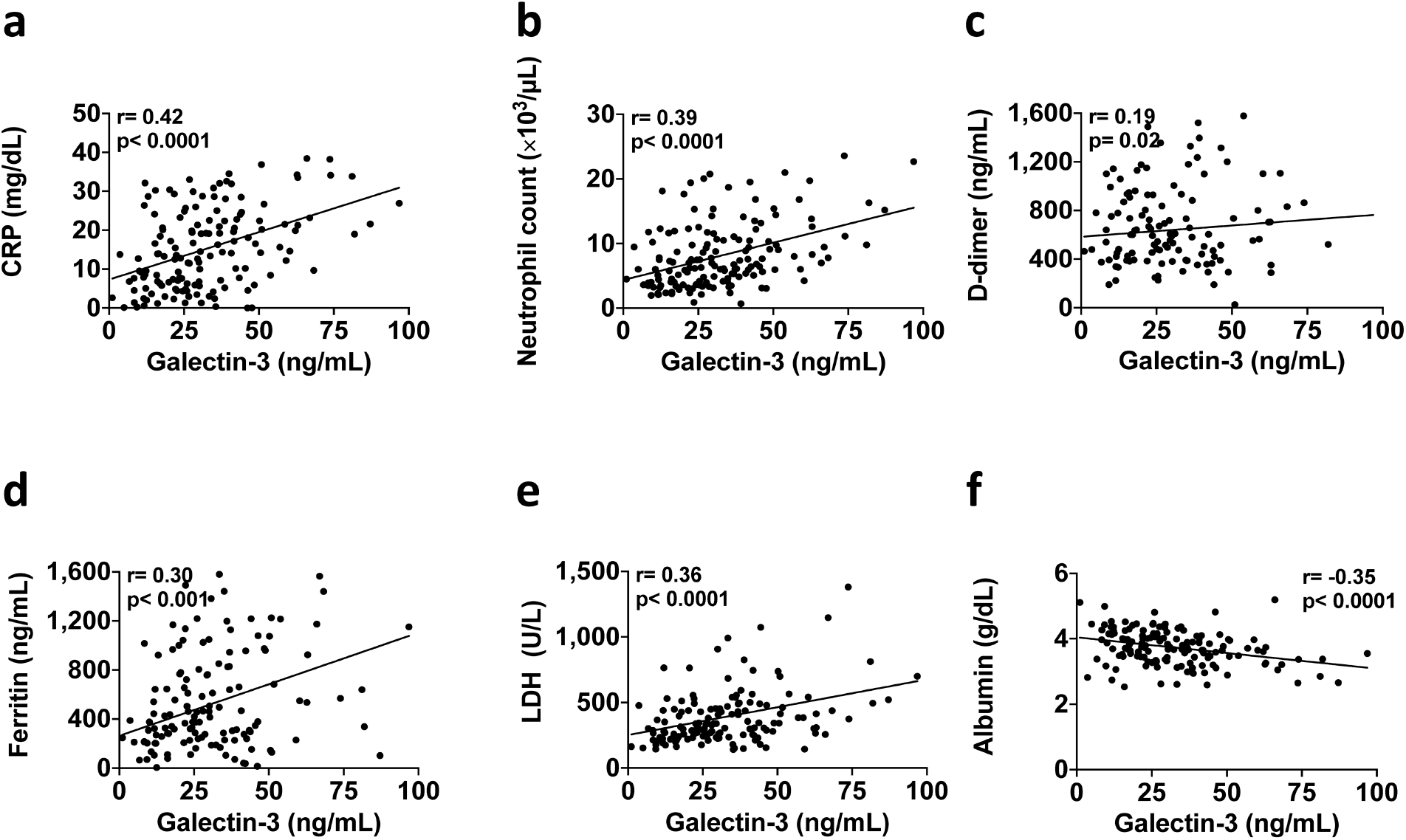
Galectin-3 correlates with inflammatory markers in COVID-19 patients. Spearman correlations show significant associations between galectin-3 and commonly measured inflammatory markers in SARS-CoV-2 infected patients. **a**, C-reactive protein (CRP), **b**, neutrophil count, **c**, D-dimer, **d**, ferritin, **e**, Lactate dehydrogenase (LDH) and **f**, albumin.

### Galectin-3, CRP and albumin levels are independent predictors of severe outcome in COVID-19 patients

To assess the discriminative power of galectin-3 as a predictor of severe outcome, ROC curves were plotted. Galectin-3 discriminates well between critical and moderate patients with an AUC of 0.69 (95% CI 0.61 – 0.78, p<0.0001), with a cut-point of 30.59 ng/mL (57.45% sensitivity, 75.81% specificity and 2.37 likelihood ratio (LR) (**Supplementary figure 1**). Galectin-3 also discriminates well between those with severe and non-severe outcome, with an AUC of 0.75 (95% CI 0.67 – 0.84, p<0.0001), with a cut-point of 30.99 ng/mL (74.07% sensitivity, 73.53% specificity, and 2.79 LR) (**Figure 4a**).

**Fig. 4:**
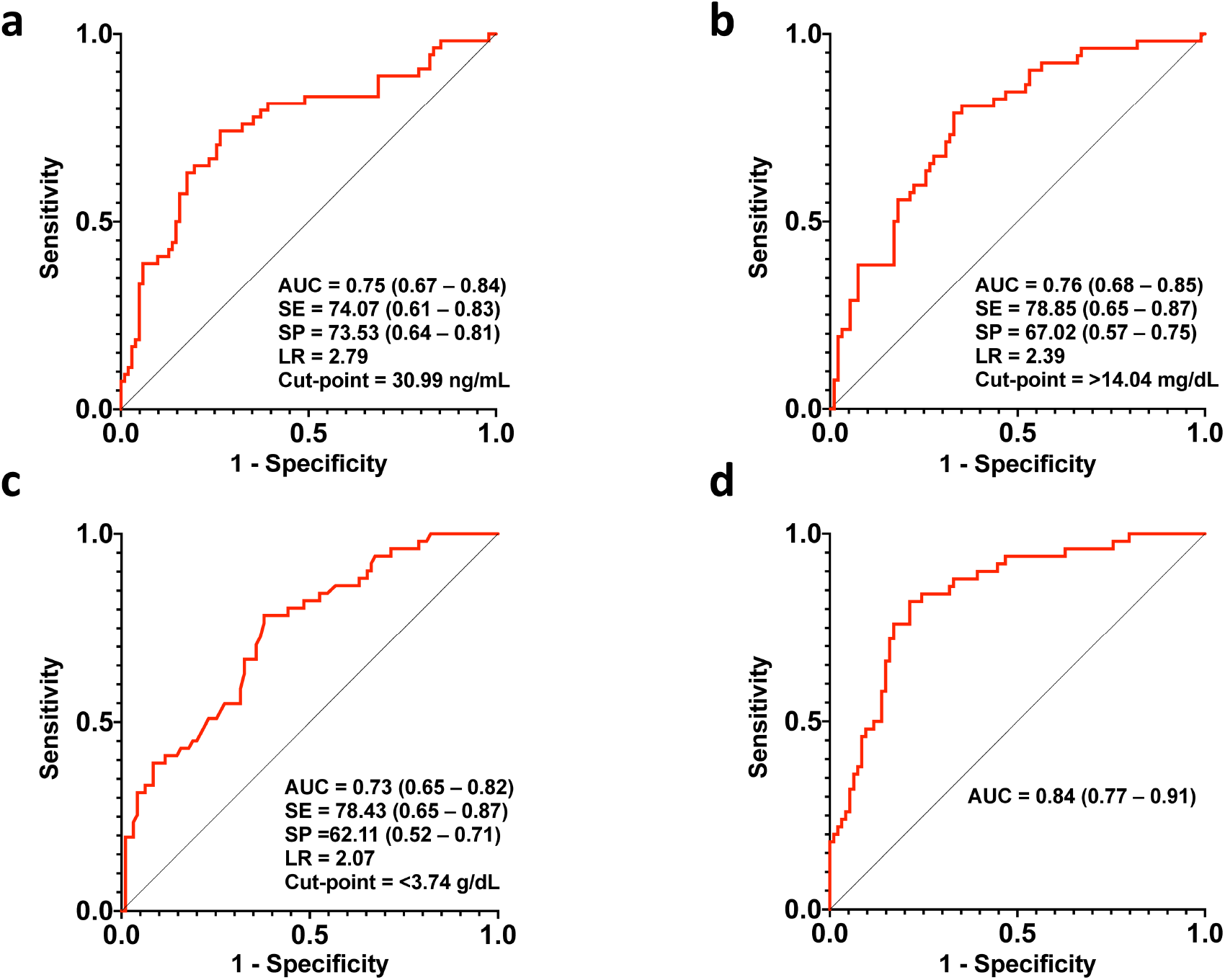
Galectin-3, albumin and CRP as independent predictors of severe outcome in COVID-19 patients. Receiver-operating characteristic curves (ROCs) of the independent predictors for the classification of binary outcomes (severe/non-severe) using **a**, galectin-3, **b**, CRP, **c**, albumin and **d**, the combined predicted probabilities of galectin-3, CRP and albumin. Values for AUC, sensitivity (SE), specificity (SP), likelihood ratio (LR) and cut-point values are shown, with 95% CIs in parentheses.

We performed a forward-stepwise logistic regression analysis to identify independent demographic and laboratory parameters that strongly correlated with a severe outcome, and thus with disease progression (i.e., IMV and/or death). A smoothing spline of galectin-3 showed a non-linear relationship with severe outcome; therefore, we used the Youden’s *J* statistic to determine the ideal binary cut-point of galectin-3 for classifying severe outcomes (**Supplementary figure 2a**). The initial model included age, gender, comorbidities and inflammatory parameters (**Supplementary table 2**). The final model selected via the stepwise procedure included galectin-3 (odds ratio [OR] 3.89 [95% CI 1.50 – 10.10], p<0.01), CRP (OR 1.05 [95% CI 1.00 – 1.11], p=0.04), and low albumin levels upon admission (OR 0.24 [95% CI 0.09 – 0.65], p<0.01) when regressed on severe outcome (IMV and/or death; **Table 2**). Of note, CRP and albumin were entered as continuous variables according to their smoothing splines which showed linear relationships with severe outcome (**Supplementary figure 2b – c)**. The equation fitted by this logistic regression model was the following, where *p = P*(*Y =* 1): 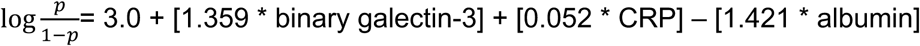, where galectin-3 is coded as binary (less than 30.99 ng/mL with 0 and above 30.99 ng/mL with 1), and CRP and albumin as continuous variables. The obtained values were transformed into predicted probabilities with the formula 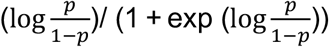.

**Table 2.** Odds ratios from the multivariable logistic regression model for severe outcome.

| **Independent predictor** | **Odds Ratio** | **95% CI** | ***p* value** |
| --- | --- | --- | --- |
| Galectin-3 (binary) | 3.89 | (1.50 – 10.10) | **<0.01** |
| CRP | 1.05 | (1.00 – 1.11) | **0.04** |
| Albumin | 0.24 | (0.09 – 0.65) | **<0.01** |

Independent predictors for severe outcome. Galectin-3 was entered as a binary variable according to its non-linear relationship with severe outcomes (>30.99 ng/mL=1, <30.99 ng/mL=0), C-reactive protein (CRP) and albumin were entered as continuous variables. Additional variables included in the model were: gender, age, diabetes, hypertension, obesity (BMI ≥30), INR, lymphocyte and neutrophil count, NLR, triglycerides. Bold values represent $p < 0.05$ .

We also explored the potential for using binary cut-points of CRP and albumin to classify severe outcomes. CRP had an AUC of 0.76 (95% CI 0.68 – 0.85, p<0.0001) at a cut-point of 14.04 mg/dL (78.85% sensitivity, 67.02% specificity and 2.39 LR), while albumin had an AUC of 0.73 (95% CI 0.65 – 0.82, p<0.0001) with a 3.74 g/dL cut-point (78.43% sensitivity, 62.11% specificity and 2.07 LR) (**Figure 4b – c**). We then assessed if the combination of galectin-3, CRP and albumin could better classify severe outcomes in COVID-19 patients than either marker on its own. To determine this, the predicted probabilities for this combination of values were computed and plotted in a ROC curve (**Figure 4d**). The AUC showed an enhanced ability to classify severe outcomes compared to each value independently 0.84 (95% CI 0.77 – 0.91, p<0.0001).

## DISCUSSION

In this prospective cohort of COVID-19 patients, we assessed the classification performance of circulating galectin-3 levels obtained upon hospitalization on the development of a severe outcome, defined as requirement of IMV and/or death. We hypothesized that this molecule could be associated with symptom severity due to its known involvement in the exacerbated inflammatory response, a feature that has been exhibited in COVID-19 patients (3).

The hyperinflammatory state in COVID-19 patients and its relationship with galectin-3 was observed in this study, as higher levels of this lectin upon admission were found in critical patients with lung affection greater than 50%. Our results indicate that galectin-3 levels above 30.59 ng/mL discriminate between patients with critical or moderate disease with a high specificity. ARDS is characterized by a diffuse alveolar damage in the lung, caused by the severe inflammatory process (25). The high levels of galectin-3 found in these patients might be an indication of the possible role of galectin-3 in the pathophysiology of ARDS and might be a reflex of the excessive inflammatory response associated with ARDS in these patients.

Evidence in the literature has implicated the cytokine release syndrome as the main factor responsible for the high mortality observed in COVID-19 patients (26). Specifically, IL-6 and TNF-α have been found to be independent predictors of severity and poor outcome (3). Galectin-3 is a leading orchestrator of the inflammatory response syndrome by activating and triggering the release of inflammatory cytokines. Our observations indicate that higher galectin-3 levels are found in those patients with a severe outcome. Furthermore, we found that galectin-3 possesses power as an independent predictor of severe outcome when adjusting for age, gender, comorbidities and other inflammatory parameters. ARDS in COVID-19 leads to more severe outcomes than ARDS due to other causes (27). With a general mortality of 26%-61.5% in those admitted to the intensive care unit, and significantly higher in those requiring IMV (65.7% to 94%) (27). Our results indicate that values greater than 30.99 ng/mL have a high sensitivity and specificity to predict an adverse clinical course with the possibility of requiring IMV and/or death. Galectin-3 was not only able to classify a severe outcome in critical patients, but, more importantly, was able to identify severe outcomes in moderate patients (AUC= 0.95).

While IMV is intended to minimize the progression of lung injury (28), it has been also demonstrated to induce or aggravate lung damage and in the long-run may contribute to lung fibrosis (4, 29). Chronic pulmonary fibrosis has been observed in recovered COVID-19 patients (10, 24). Galectin-3 is known to play a role in the pathogenesis of pulmonary fibrosis, and clinical trials testing galectin-3 inhibitors are currently underway for the treatment of idiopathic pulmonary fibrosis (30). Galectin-3 could provide an important biomarker for severe COVID-19 with potential for involvement in the direct pathophysiological process of the underlying disease.

To better understand the relationship between galectin-3 and the inflammatory response after SARS-CoV-2 infection, we explored its association with CRP. CRP, an acute inflammatory biomarker with ability to predict mortality in COVID-19 (31, 32), was identified as an independent predictor of severe outcome in our cohort and had a positive correlation with galectin-3. This novel association between galectin-3 and CRP has not been reported in viral infection, much less in COVID-19 but it suggests the utility of this molecule in detecting the inflammatory state of patients upon hospital arrival. As both CRP and galectin-3 were identified as independent predictors, we sought to identify which one would perform better according to its association with patient outcome. The non-linear relationship of galectin-3, observed in a smoothing spline, demonstrated that higher levels of this lectin were a common characteristic of patients at high-risk of progressing to a severe COVID-19 outcome. We consider that inflammatory markers such as CRP which tend to show a linear relationship with outcome may not be suitable as efficient biomarkers given their inconsistency, as different cut-off values have been reported; therefore galectin-3 may present an advantage over this commonly measured laboratory parameter, as it showed more accuracy when identifying patients prone to disease progression. Furthermore, although a slope could be interpreted by healthcare providers for continuous predictors, we also appreciated the simplicity of using a binary cut-point of galectin-3 level to classify patients as low or high risk for severe outcomes.

Part of the relevance of this molecule in COVID-19 may be also explained given its interaction with neutrophils, a crucial immune cell whose recruitment is essential in the innate immune response against invading pathogens (e.g., SARS-CoV-2). Neutrophilia is also believed to be a key aspect in the cytokine release that has been studied in severe patients (33) which in this cohort was positively correlated with galectin-3, consequently supporting its role in the acute pro-inflammatory response. Moreover, a previous report has shown that galectin-3 is an adhesion molecule that mediates neutrophil adhesion to endothelial cells during *Streptococcus pneumoniae* infection, confirming its association with lung inflammation as it is actively released after infection (34).

In accordance with earlier studies (35), we also identified hypoalbuminemia as a common characteristic among critically ill patients and support the correlation between albumin and the systemic inflammation in COVID-19 patients, as a negative correlation was found with galectin-3, which is a known contributor to the release of pro-inflammatory cytokines (12). Albumin is an important biomarker that reflects the inflammatory state, as its production is decreased due to higher levels of IL-6 (36). Observations carried out by Huang et al. in a large cohort of COVID-19 patients identified the decrease in albumin levels as a significant indicator of progression to a critical stage and death. They associated this pathologic finding with a reduced capacity of synthesis by hepatocytes as mild hepatic injury was evident (37). Another aspect relevant to consider is that capillary leakage into the interstitial space increases in severe illness such as sepsis, leading to the sequestration of albumin (38).

As galectin-3 was found to reflect the hyperinflammatory state of patients, its predictive ability together with CRP and albumin was tested. Results revealed that when used jointly, severe outcomes can be more accurately classified upon hospital admission (AUC=0.84), thus providing clinicians more resources to efficiently identify patients with higher odds of adverse progression. Given that galectin-3 is highly correlated with the severity of the disease, as shown when patients were first classified based of CT findings, we propose that galectin-3 be included in the initial screening of COVID-19 patients, together with CRP and albumin. The assessment of this panel upon admission will help identify patients at high-risk of disease progression as galectin-3 was found to be associated with the patients’ clinical state. Therapies that decrease inflammatory response would be expected to reduce galectin-3 levels during the clinical course of the patient and influence outcomes. Further studies are needed to assess whether galectin-3 can provide a useful biomarker to evaluate therapeutic interventions.

In this study, we have offered evidence on the prognostic use of galectin-3 in SARS-CoV-2 infected patients which may extend to other critical diseases and propose its combined use with other inflammatory markers to guide the clinical rationale when assessing a hospitalized patient’s risk. Inhibitors of galectin-3 have been shown to reduce the levels of both IL-6 and TNF-α in vitro and have shown anti-inflammatory effects *in vivo* (39). Based on the data presented here, we also propose galectin-3 as a feasible pharmacological target to minimize the hyperinflammatory phase and the subsequent lung fibrosis in COVID-19 patients.

There are some limitations to our study. First, since this is a single-center experience, data from different populations and a multicenter analysis will be needed for validation. Second, due to the small sample size, further clinical studies with larger sample sizes are required to confirm these findings before galectin-3 can definitively be recommended as a standard biomarker in the hospital setting. Despite these limitations, this study demonstrates in a prospective cohort of COVID-19 patients at one of the largest health institutes in Mexico that measurement of galectin-3 levels upon hospital admission could be helpful in predicting disease severity. Finally, the combined use of galectin-3, CRP and albumin showed strong predictive ability, and thus could aid to efficiently allocate medical resources before patients develop an adverse outcome.

## METHODS

### Study design and groups

156 patients admitted to the Instituto Nacional de Ciencias Médicas y Nutrición Salvador Zubirán (INCMNSZ), one of the largest designated institutions in Mexico for the hospitalization of patients with COVID-19 were prospectively included according to the following criteria: laboratory confirmed COVID-19 by RT-PCR, CT and clinical characteristics consistent with COVID-19. Those who presented with respiratory symptoms but had a negative RT-PCR test result were excluded from the analyses. Patients were divided into two groups based on the pulmonary affection on a CT. Moderate patients: those with <50% of lung damage. Critical patients: those with >50% of lung damage. This study was approved by INCMNSZ’s Research Ethics Committee (No. GAS-3385-20-20-1) and complied with the provisions of the Declaration of Helsinki. Informed written consent was obtained from all patients prior to blood sample collection.

### Primary outcome definition

Patients who required IMV and/or died during hospitalization were categorized as having a *severe outcome*. Patients who recovered and were discharged without requiring IMV were categorized as having a *non-severe outcome*.

### Data collection

Clinical and laboratory data were extracted from the electronic medical records including: Demographics (age, gender, comorbidities), clinical (days of hospital stay), radiological (chest CT findings), laboratory and patient outcome data (need for IMV and/or death). Laboratory data included arterial blood gas, complete blood count, triglycerides, albumin, lactate dehydrogenase, liver enzymes, coagulation tests (D-dimer, INR) and inflammation-related parameters (CRP and ferritin).

### Sample collection and Galectin-3 levels measurement

Blood samples were collected upon hospital admission from all 156 patients. Samples were centrifuged at 3,000 rpm for 10 min, and serum was aliquoted and stored at −70°C until further analysis. Galectin-3 was measured in the serum samples using a commercial ELISA Kit (Invitrogen, #BMS 279-4, Carlsbad, CA, USA), according to the manufacturing instructions. All samples were evaluated in duplicate. The inter-assay coefficient of variation was 8.52% and the intra-assay 5.34%.

### Statistical analysis

Data are expressed as frequencies for categorical variables and as mean with standard deviation (SD) or median with interquartile range (IQR) for continuous variables according to their distributions. Student’s t-tests or Mann-Whitney U tests were used for univariate statistical comparisons, while correlation analyses were performed with Spearman’s correlation coefficient for pairs of continuous variables. To determine the prognostic ability of galectin-3 and inflammatory markers for the primary outcome, receiver operating characteristic (ROC) curves were plotted, and cut-point values were chosen as those with the highest Youden’s *J* statistic. Independent predictors of the primary outcome were determined using multivariable logistic regression analysis (forward-stepwise selection). This analysis included variables with a p value <0.20 in bivariate analyses; goodness of the fit was assessed with the Hosmer-Lemeshow test. The combined power of the identified independent predictors was evaluated with a ROC curve using the model selected by the stepwise logistic regression procedure. Statistical analyses were performed with SPSS (version 24.0, SPSS Inc., Chicago, IL, USA) and GraphPad Prism (version 8.00, GraphPad Software, La Jolla, CA, USA). A selected alpha level of 0.05 indicated statistical significance.

## Data Availability

The authors confirm that the data supporting the findings of this study are available within the article [and/or] its supplementary materials

## AUTHOR CONTRIBUTIONS

N.L.R., E.C.A. and N.N.A. conceived and designed the study. N.L.R, E.C.A, M.S.M., P.V.S., M.P.J., F.R.A., B.I.V.M., E.A.A, J.M.E.V., J.A., O.M.G., F.T.D., J.T.R., D.G.M., and N.N.A., collected clinical data. N.L.R, E.C.A. and N.N.A. contributed to data analysis. K.L.C. and O.M.G provided input on statistical methods. N.L.R and E.C.A. wrote the manuscript and generated tables and figures. N.N.A., C.A.H., D.K. and K.L.C. contributed to the manuscript review. All authors critically reviewed and approved the final version of the manuscript.

## COMPETING INTERESTS

The authors declare no conflicts of interest that pertain to this work

## FINANCIAL SUPPORT STATEMENT

This work was supported by the grant #312285 from Consejo Nacional de Ciencia y Tecnología (CONACYT)-México (N.N.A.) and University of Colorado Anschutz Medical Campus Department of Surgery Academic Enrichment Fund (C.A.H.)

## ABBREVIATIONS

ARDS: Acute respiratory distress syndrome
COVID-19: Coronavirus disease 2019
CRP: C-Reactive protein
CT: Computerized tomography
ELISA: Enzyme-linked immunosorbent assay
IMV: Invasive mechanical ventilation
RT-PCR: Real-time reverse transcriptase-polymerase chain reaction
SARS-CoV-2: Severe acute respiratory syndrome coronavirus-2

---

**Supplementary fig. 1:**
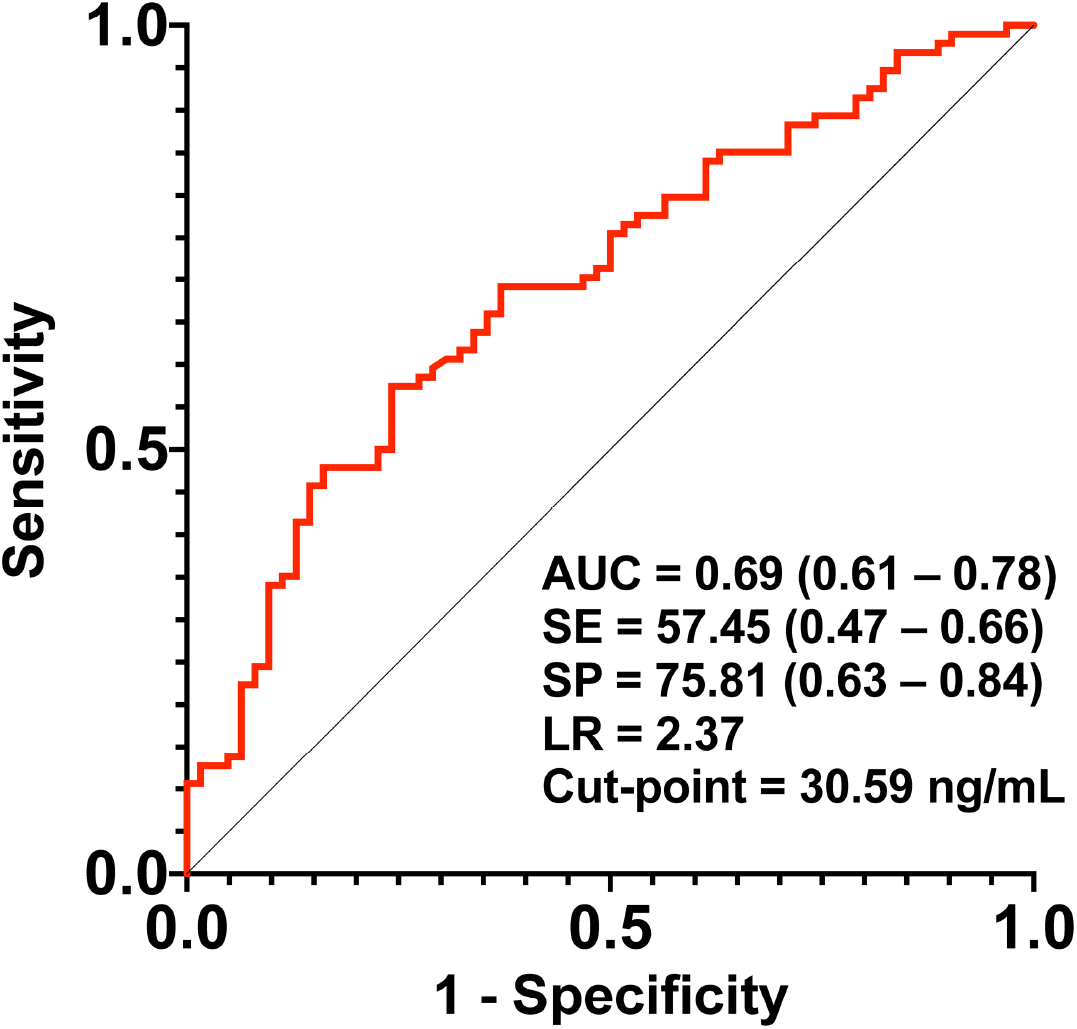
Galectin-3 discriminates between critical and moderate patients. Galectin-3 associates with CT findings upon hospital admission as it shows power discriminating between critical patients (>50% of lung affection) and moderate patients (<50% of lung affection). Values for AUC, sensitivity (SE), specificity (SP), likelihood ratio (LR) and cut-point values are shown, with 95% CIs in parentheses.

**Supplementary fig. 2:**
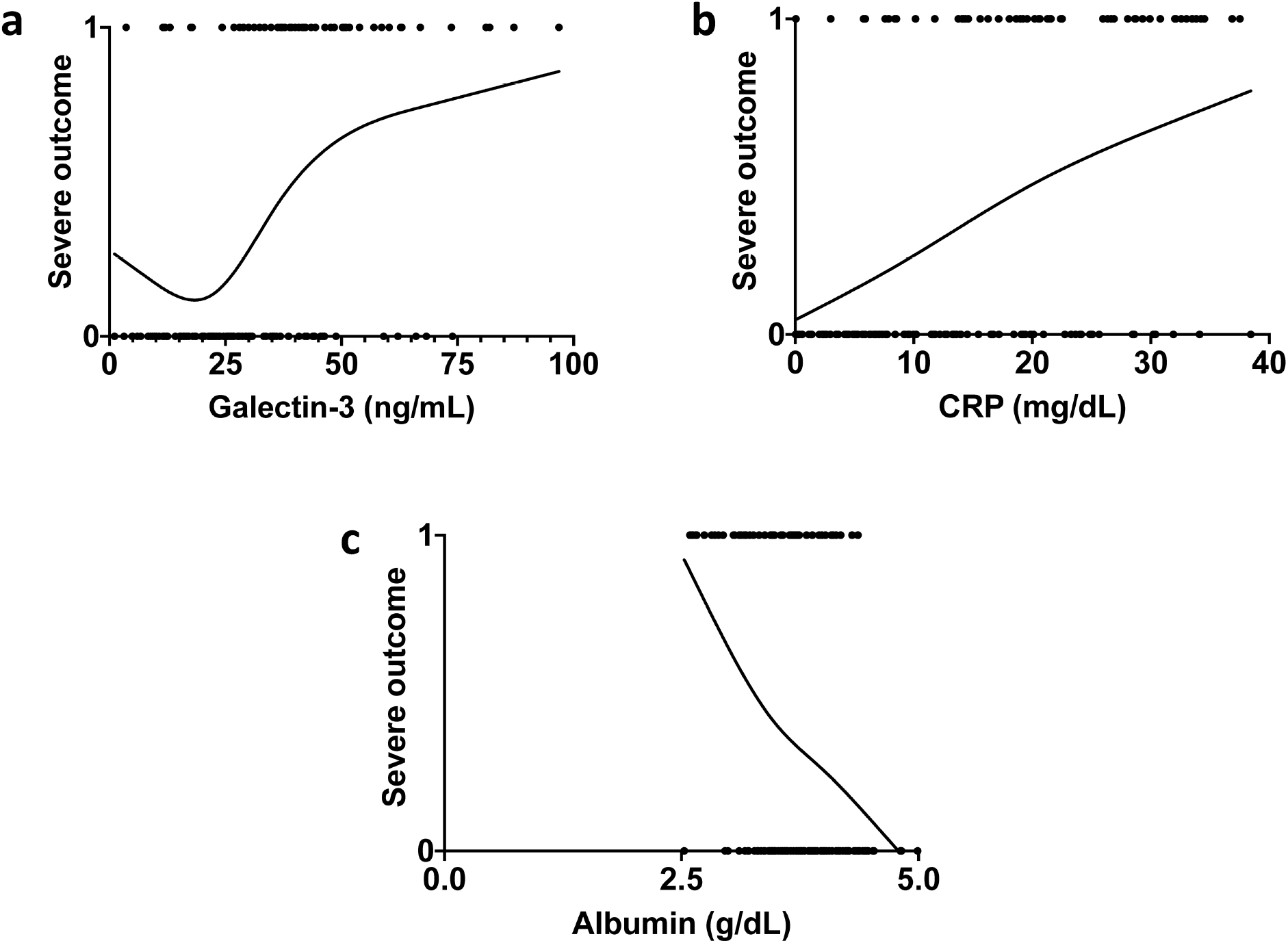
Smoothing splines of galectin-3, CRP and albumin. **a**, Galectin-3 shows a non-linear relationship with the patients’ outcome (severe=1 and non-severe=0), for which it is used as a binary variable in the logistic regression analysis. **b**,**c**, Smoothing splines of CRP (**b**) and albumin (**c**), showing a linear relationship with outcome, using both as continuous variables in further analysis. Data in **a**,**b**,**c** are computed with 4 knots.

## Notes

### Competing Interest Statement

The authors have declared no competing interest.

### Author Declarations

This study was approved by the Instituto Nacional de Ciencias Médicas y Nutrición Salvador Zubirán's Research Ethics Committee (No. GAS-3385-20-20-1).

