## Supplementary Table 1 and 2 for "Galectin-3 as a potential prognostic biomarker of severe COVID-19 in SARS-CoV-2 infected patients"

**Supplementary Table 1. Demographic characteristics, healthy subjects.**

| | **Total (n= 10)** |
|---|---|
| Age | 48.70 ± 5.48 |
| Gender, female | 6 (60.0%) |
| Weight | 59.12 ± 6.34 |
| BMI | 22.81 ± 1.35 |
| Hypertension | 0 (0.0%) |
| Diabetes | 0 (0.0%) |
| Alcohol consumption | 2 (20.0%) |
| Galectin-3 (ng/mL) | 9.65 [8.27 – 14.71] |
| Glucose (mg/dL) | 89.8 ± 6.07 |
| Triglycerides (mg/dL) | 111.70 ± 46.31 |
| Creatinine (mg/dL) | 0.78 ± 0.14 |
| ALT (U/L) | 24.0 ± 8.47 |
| AST (U/L) | 25.6 ± 4.40 |

Data are reported as mean (± SD), median (IQR) and *n* (%). BMI= Body mass index, ALT= alanine transaminase, AST= aspartate transaminase.

**Supplementary Table 2. Forward-stepwise multivariable logistic regression**

| | **Variables** | ***p* value** |
|---|---|---|
| **Step 0** | Age | 0.260 |
| | Gender, male | 0.491 |
| | Diabetes | 0.244 |
| | Hypertension | 0.317 |
| | Galectin-3 (binary) | < 0.0001 |
| | INR | 0.047 |
| | Albumin | < 0.0001 |
| | Triglycerides | < 0.01 |
| | Ferritin | 0.01 |
| | D-Dimer | < 0.01 |
| | CRP | < 0.0001 |
| | Lymphocyte count | 0.334 |
| | Neutrophil count | <0.001 |
| | NLR | <0.01 |

| | **Variables** | **OR (95% CI)** | ***p* value** |
|---|---|---|---|
| **Step 1** | Galectin-3 (binary) | 6.65 (2.82 – 15.70) | < 0.0001 |
| **Step 2** | Galectin-3 (binary) | 5.00 (2.00 – 12.45) | < 0.001 |
| | Albumin | 0.19 (0.07 – 0.51) | < 0.01 |
| **Step 3** | Galectin-3 (binary) | 3.89 (1.50 – 10.10) | < 0.01 |

| | | | |
|---|---|---|---|
| | CRP | 1.05 (1.00 – 1.11) | 0.04 |
| | Albumin | 0.24 (0.09 – 0.65) | < 0.01 |

Variables included in every step of the forward-stepwise logistic regression. INR= International normalized ratio, CRP= C-reactive protein, NLR= Neutrophil to lymphocyte ratio.
